# Comparison of prehospital assessment by paramedics and in-hospital assessment by physicians in suspected stroke patients

**DOI:** 10.1101/2023.05.08.23289702

**Authors:** Luuk Dekker, Jasper D. Daems, Martijne H.C. Duvekot, T. Truc My Nguyen, Esmee Venema, Adriaan C.G.M. van Es, Anouk D. Rozeman, Walid Moudrous, Kirsten R.I.S. Dorresteijn, Jan-Hein J. Hensen, Jan Bosch, Erik W. van Zwet, Els L.L.M. de Schryver, Loet M.H. Kloos, Karlijn F. de Laat, Leo A.M. Aerden, Ido R. van den Wijngaard, Diederik W.J. Dippel, Henk Kerkhoff, Marieke J.H. Wermer, Bob Roozenbeek, Nyika D. Kruyt, the PRESTO and LPSS investigators

**Author notes:** Corresponding author: Luuk Dekker, MD, Department of Neurology, Leiden University Medical Center, Albinusdreef 2, P.O. Box 9600, 2300 RC, Leiden, the Netherlands. both authors contributed equally to the manuscript.

## Abstract

**Background:** It is unknown if ambulance paramedics adequately assess neurological deficits used for prehospital stroke scales to detect anterior large-vessel occlusions. We aimed to compare prehospital assessment of these stroke-related deficits by paramedics with in-hospital assessment by physicians.

**Methods:** We used data from two prospective cohort studies: the Leiden Prehospital Stroke Study (LPSS) and PREhospital triage of patients with suspected STrOke (PRESTO) study. In both studies, paramedics scored 9 neurological deficits in stroke code patients in the field. Trained physicians scored the National Institutes of Health Stroke Scale (NIHSS) at hospital presentation. Patients with transient ischemic attack were excluded because of the transient nature of symptoms. Spearman’s rank correlation coefficient (r_s_) was used to assess correlation between the total prehospital assessment score, defined as the sum of all prehospital items, and the total NIHSS score. Correlation, sensitivity and specificity were calculated for each prehospital item with the corresponding NIHSS item as reference.

**Results:** We included 2850 stroke code patients. Of these, 1528 had ischemic stroke, 243 intracranial hemorrhage, and 1079 stroke mimics. Correlation between the total prehospital assessment score and NIHSS score was strong (r_s_=0.70; 95%CI: 0.68-0.72). Concerning individual items, prehospital assessment of arm (r_s_=0.68) and leg (r_s_=0.64) motor function correlated strongest with corresponding NIHSS items, and had highest sensitivity (arm 95%, leg 93%) and moderate specificity (arm 71%, leg 70%). Neglect (r_s_=0.31), abnormal speech (r_s_=0.50) and gaze deviation (r_s_=0.51) had weakest correlations. Neglect and gaze deviation had lowest sensitivity (52% and 66%) but high specificity (84% and 89%), whilst abnormal speech had high sensitivity (85%) but lowest specificity (65%).

**Conclusions:** The overall prehospital assessment of stroke code patients correlates strongly with in-hospital assessment. Prehospital assessment of neglect, abnormal speech and gaze deviation differed most from in-hospital assessment. Focused training on these deficits may improve prehospital triage.

## INTRODUCTION

In order to prevent delays in treatment of patients with ischemic stroke due to anterior circulation large vessel occlusion (aLVO), the AHA/ASA stroke triage algorithm recommends the use of prehospital aLVO stroke scales to help ambulance paramedics identify aLVO patients for direct transportation to a thrombectomy-capable stroke center (TSC).^1^ However, sensitivity of these scales is relatively low, ranging between 38% and 67%.^2,3^ A possible explanation for this is that neurological deficits are not adequately recognized by ambulance paramedics in the field. Ambulance paramedics only encounter stroke code patients sporadically during their shifts and adequate assessment of neurological deficits could require more extensive training. Meanwhile, neurological deficits are routinely assessed with the standardized National Institute of Health Stroke Scale (NIHSS) by experienced physicians upon arrival at the hospital.^4^ Some studies have compared the overall prehospital assessment of aLVO stroke scales by paramedics with assessment of physicians, but focused on total scores of a single scale and did not evaluate separate items.^5-8^ Only four small studies have evaluated to what extent examination of individual neurological deficits by paramedics matches the in-hospital assessment by physicians.^9-12^ Furthermore, only two of these studies included all stroke code patients, including patients with a stroke mimic, and in only one the paramedical assessment was performed on-site in the prehospital setting.^9,10^ Further knowledge of the level of agreement could guide training programs for paramedics and improve prehospital aLVO triage by focusing training on the more challenging neurological deficits. Therefore, we aimed to compare prehospital assessment of stroke code patients by paramedics with in-hospital assessment by physicians.

## METHODS

### Patients

We used data from stroke code patients included in the Leiden Prehospital Stroke Study (LPSS) and the PREhospital triage of patients with suspected STrOke (PRESTO) study.^2,3^ LPSS and PRESTO were both prospective multiregional observational cohort studies that included stroke code patients from four ambulance regions in the Netherlands. Together these regions serve roughly 3.7 million inhabitants with 10 PSCs and 5 TSCs, including 13 ambulance posts with approximately 600 paramedics. Both studies included all patients aged ≥18 years in whom an acute stroke code was activated by ambulance paramedics. A stroke code was activated in case of a positive Face-Arm-Speech-Time (FAST) test (PRESTO) or other acute neurological deficits at the insight of the individual paramedic (LPSS). The final diagnosis was made by local clinicians either at discharge (PRESTO) or after 3 months (LPSS). In PRESTO an imaging core laboratory committee re-assessed CT angiographies for presence of aLVO. The complete study designs, procedures and results of the PRESTO and LPSS studies can be found elsewhere.^2,3,13^ For this study, we excluded patients with a final diagnosis of transient ischemic attack (TIA) since the transient nature of symptoms would hamper a fair comparison. We used the STROBE guidelines for reporting this observational study.^14^

### Prehospital and in-hospital assessment

Paramedics scored 9-11 neurological deficits from the NIHSS on-site upon arrival at the patient and stored these in a web-based application (**Table S1**). In PRESTO, the following nine items were assessed: answering questions, following commands, abnormal speech, gaze deviation, facial palsy, arm motor function, leg motor function, grip strength, and agnosia (recognition of own arm and/or impairment). In the LPSS, the same items were assessed with the addition of sensory deficits and tactile extinction. Completing all items was required in PRESTO. In the LPSS paramedics could score the items abnormal speech, grip strength, arm and leg motor function, and tactile extinction as ‘untestable’, e.g. if the patient had a decreased level of consciousness or was fixed for transport. For this study, all untestable items were scored as missing. In the Netherlands, ambulance paramedics are registered nurses, who generally completed an additional 2-year training in either intensive, cardiac, anesthetic or emergency medical care, plus a one-year training in ambulance care. Prior to the initiation of each study, they received a brief introductory session (30-45 minutes) focusing on the use of the application and assessment of the clinical items required for the studied prehospital aLVO scales. At the emergency department, the NIHSS was assessed as part of routine work-up by a trained neurologist, neurology resident or emergency physician from the stroke team. In case NIHSS items were not documented these were reconstructed using a standardized scoring chart (PRESTO) or validated algorithm (LPSS) based on neurological examination reports.^15^ The time between ambulance arrival at the patient location and hospital arrival (ambulance-arrival-to-door time) was used to estimate the time-interval between the prehospital and in-hospital assessment.

### Statistical analysis

We analyzed the correlation between the total prehospital assessment score (range 0-15) and the total in-hospital NIHSS score (range 0-42) using Spearman’s rank correlation coefficient (r_s_) and constructed 95% confidence intervals (CIs) using Fisher’s Z transformation. The total prehospital assessment score was defined as the summation of all scores of the 9 stroke items that were assessed in both the LPSS and PRESTO. Patients with incomplete prehospital or NIHSS assessments were excluded from this analysis. To evaluate possible differences in assessment between diagnoses, this analysis was repeated after stratifying by final diagnosis.

Similarly, since spontaneous clinical improvement or deterioration might occur especially in the first few hours after onset, we also assessed Spearman’s correlation after dividing the cohort in quartiles based upon the time between onset and ambulance arrival.^16-18^ A Spearman’s coefficient of 0-0.19 was regarded as very weak, 0.2-0.39 as weak, 0.40-0.59 as moderate, 0.6-0.79 as strong and 0.8-1 as very strong correlation.^19^

Second, to identify more challenging items, we calculated the correlation, sensitivity and specificity of each prehospital item compared to its corresponding in-hospital item from the NIHSS. To calculate sensitivity and specificity, we dichotomized all prehospital and in-hospital findings as either normal or abnormal, using the in-hospital NIHSS as reference. We constructed 95% CIs for sensitivity and specificity using the normal approximation method based on estimated standard errors. If LPSS and PRESTO used different scoring methods for the same item, scores were combined and adjusted to match the NIHSS (**Table S1**). For correlation, the combined score of LPSS and PRESTO was compared with the NIHSS. Because prehospital assessment of arm motor function did not include an isolated drift without paresis in PRESTO, a drift on the LPSS and NIHSS item arm motor function was scored as normal. Similarly, since both PRESTO and LPSS did not score a drift without paresis for leg motor function, a drift on this NIHSS item was also scored as normal. The prehospital item abnormal speech was compared to any symptoms on the language or dysarthria items on the NIHSS. Prehospital assessment of agnosia was compared with the NIHSS item neglect. In PRESTO, agnosia was only scored in patients with left sided hemiparesis, according to the Rapid Arterial oCclusion Evaluation (RACE) scale.^20^ Additionally, because tactile extinction was also assessed in the LPSS and since the NIHSS item neglect encompasses more modalities than only agnosia, we also compared neglect with the combined prehospital assessment of agnosia and tactile extinction in the LPSS cohort. Patients with missing prehospital or NIHSS assessments were excluded on a per item basis for these comparisons. Lastly, grip strength was excluded from the analysis of individual items, since there is no corresponding item on the NIHSS.

Statistical analyses were performed using R (version 4.1.2) and R Studio (version 2022.2.0.443).

## RESULTS

Of the 3320 included stroke code patients, 470 had a TIA and were excluded (**Figure 1**). Of the remaining 2850 patients, 1528 (53.6%) had an ischemic stroke (295 with aLVO), 243 (8.5%) had an intracranial hemorrhage and 1079 (37.9%) a stroke mimic. Median age was 73 years and 49% were female (**Table 1**). Median ambulance-arrival-to-door time was 29 minutes (interquartile range (IQR) 23-36 minutes). Median total prehospital assessment score was 2 and in-hospital NIHSS score was 3. These were higher for aLVO ischemic stroke patients (median prehospital assessment score: 8, NIHSS score: 12) and patients with an intracranial hemorrhage (prehospital assessment score: 6, NIHSS score: 10) (**Table S2**).

**Figure 1.**
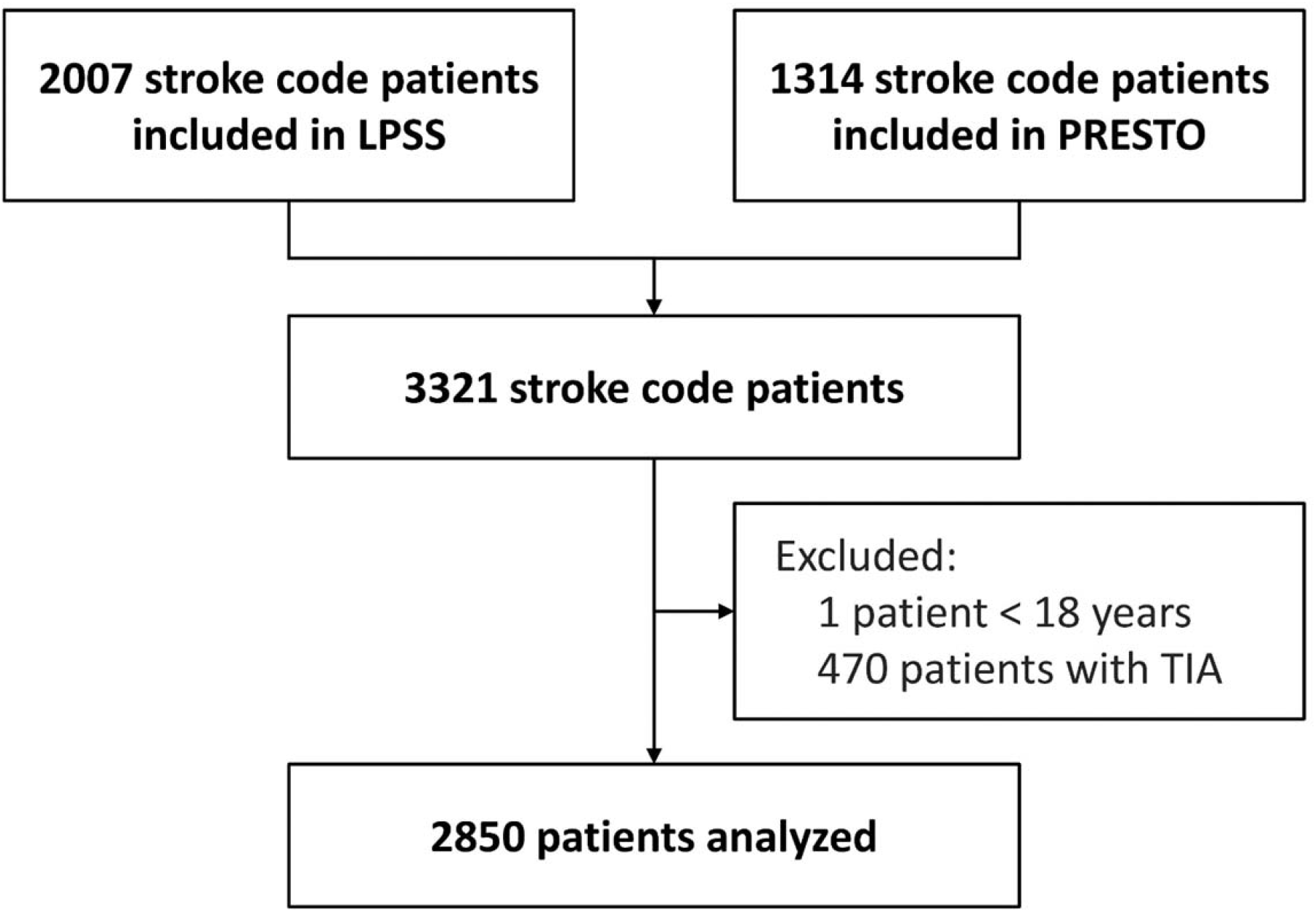
Flowchart of analyzed patients. *LPSS = Leiden Prehospital Stroke Study; PRESTO = PREhospital triage of patients with suspected STrOke study; TIA = transient ischemic attack*

**Table 1.** Baseline characteristics of analyzed patients.

| Characteristic | n=2850 |
| --- | --- |
| Age, median (IQR) | 73 (61-82) |
| Female sex, n/N (%) | 1401/2850 (49) |
| Premorbid mRS 0-2, n/N (%) | 2208/2641 (84) |
| History of: |  |
| atrial fibrillation, n/N (%) | 421/2829 (15) |
| hypertension, n/N (%) | 1720/2835 (61) |
| hypercholesterolemia, n/N (%) | 1470/2828 (52) |
| diabetes mellitus, n/N (%) | 579/2830 (21) |
| ischemic stroke or TIA*, n/N (%) | 811/2832 (29) |
| myocardial infarction, n/N (%) | 316/2825 (11) |
| intracranial hemorrhage, n/N (%) | 86/2841 (3) |
| Use of antiplatelet medication, n/N (%) | 1002/2822 (36) |
| Use of oral anticoagulation, n/N (%) | 457/2809 (16) |
| Primarily presented at a PSC, n/N (%) | 1196/2850 (42) |
| Primarily presented at a TSC, n/N (%) | 1654/2850 (58) |
| Onset-to-ambulance-arrival time, in minutes, median (IQR) | 91 (30-277) |
| Ambulance-arrival-to-door time, in minutes, median (IQR) | 29 (23-36) |
| Total prehospital assessment score (range 0-15), median (IQR) | 2 (1-5) |
| Total in-hospital NIHSS score (range 0-42), median (IQR) | 3 (1-7) |

\* In PRESTO only information on a history of ischemic stroke was collected.
*IQR = interquartile range; mRS = modified Rankin Scale; NIHSS = National Institutes of Health Stroke Scale; PSC = primary stroke center; TIA = transient ischemic attack; TSC = thrombectomy-capable stroke center*

Spearman’s correlation coefficient between the total prehospital assessment score and in-hospital NIHSS score was strong (0.70; 95% CI: 0.68-0.72) (**Table 2, Figure S1**). After stratification by final diagnosis, correlation was moderate for patients with a stroke mimic (0.55, 95% CI: 0.49-0.59), strong for patients with non-aLVO ischemic stroke (0.61, 95% CI: 0.56-0.64), stronger for aLVO ischemic stroke (0.76, 95% CI: 0.69-0.81), and very strong for intracranial hemorrhage (0.83, 95% CI: 0.77-0.87) (**Table 2**). As can be seen in **Table S3**, the correlation was highest in the first quartile, but fluctuated throughout the entire onset-to-ambulance-arrival time range.

**Table 2.** Correlation between the total prehospital assessment score (range 0-15) and in-hospital NIHSS score (range 0-42), and stratified by final diagnosis.

| | N* | Spearman's $r_s$<br>(95% CI) | Median total<br>prehospital<br>assessment<br>score (IQR) | Median total<br>in-hospital<br>NIHSS score<br>(IQR) |
| --- | --- | --- | --- | --- |
| **Total cohort** | 2058 | 0.70 (0.68-0.72) | 2 (1-5) | 3 (1-7) |
| **aLVO ischemic stroke** | 211 | 0.76 (0.69-0.81) | 8 (4-10) | 12 (6-17) |
| **non-aLVO ischemic stroke** | 959 | 0.61 (0.56-0.64) | 2 (1-5) | 3 (2-5) |
| **intracranial hemorrhage** | 153 | 0.83 (0.77-0.87) | 6 (3-10) | 10 (4-18) |
| **stroke mimic** | 735 | 0.55 (0.49-0.59) | 1 (0-3) | 1 (0-3) |

\* Analyses were performed in patients of whom both complete prehospital assessment and in-hospital NIHSS scores were available.
*aLVO = anterior large vessel occlusion; CI = confidence interval; IQR = interquartile range; NIHSS = National Institutes of Health Stroke Scale*

Concerning comparison of the prehospital and in-hospital items, Spearman’s coefficient was highest for arm motor function (0.68, 95% CI: 0.66-0.70) and leg motor function (0.64, 95% CI: 0.61-0.66). In contrast, correlation was poorest for neglect (0.31, 95% CI: 0.27-0.35), followed by abnormal speech (0.50, 95% CI: 0.47-0.53) and gaze deviation (0.51, 95% CI: 0.48-0.54) (**Table 3**). Sensitivity was highest for arm motor function (95%, 95% CI: 93-97) and leg motor function (93%, 95% CI: 91-96), and lowest for neglect (52%, 95% CI: 45-59) and gaze deviation (66%, 95% CI: 61-70). Whilst specificity was highest for gaze deviation (89%, 95% CI: 88-90), answering questions (87%, 95% CI: 85-88), and following commands (85%, 95% CI: 84-87), and lowest for abnormal speech (65%, 95% CI: 62-67). Furthermore, in patients with neglect on the NIHSS, the prehospital assessment of neglect was frequently missing (72/269; 27%) (**Table S4**). Also, the number of false-positive patients (n=252) was higher than true-positive patients (n=102), resulting in a very low positive predictive value (102/354 = 29%) of neglect. A complete overview of prehospital and in-hospital observations per item can be found in **Table S4**.

**Table 3.** Comparison of prehospital and in-hospital assessments of individual items.

| | N | Spearman's $r_s^*$<br>(95% CI) | Sensitivity*<br>(95% CI) | Specificity*<br>(95% CI) |
| --- | --- | --- | --- | --- |
| **Questions** | 2735 | 0.61 (0.59-0.64) | 78 (75-81) | 87 (85-88) |
| **Commands** | 2736 | 0.55 (0.52-0.57) | 74 (70-78) | 85 (84-87) |
| **Abnormal speech** | 2729 | 0.50 (0.47-0.53) | 85 (83-87) | 65 (62-67) |
| **Gaze deviation** | 2713 | 0.51 (0.48-0.54) | 66 (61-70) | 89 (88-90) |
| **Facial palsy** | 2759 | 0.58 (0.55-0.60) | 76 (73-79) | 79 (77-81) |
| **Arm motor function** | 2443 | 0.68 (0.66-0.70) | 95 (93-97) | 71 (69-73) |
| **Leg motor function** | 2225 | 0.64 (0.61-0.66) | 93 (91-96) | 70 (68-72) |
| **Neglect (compared to agnosia) <sup>†</sup>** | 1725 | 0.31 (0.27-0.35) | 52 (45-59) | 84 (82-85) |
| **Neglect (compared to agnosia and tactile extinction) <sup>‡</sup>** | 1249 | 0.26 (0.21-0.31) | 64 (54-74) | 79 (77-82) |

\* For correlation, the combined score of LPSS and PRESTO was compared with the NIHSS.
For sensitivity and specificity, items were dichotomized as either normal or abnormal and the
in-hospital NIHSS was used as reference. Values are shown in percentages (%).
<sup>†</sup> In PRESTO, agnosia was only scored in patients with left sided hemiparesis.
<sup>‡</sup> Only shown for the LPSS cohort, because tactile extinction was not examined in PRESTO.
*CI = confidence interval*

The additional analysis of the extended definition of neglect (agnosia with the addition of tactile extinction) in the LPSS cohort showed an increase in sensitivity of neglect from 52% to 64%, with a specificity of 79% (95% CI: 77-82). However, due to the low prevalence of abnormal findings (7%) and low number of complete cases (89/168), the 95% confidence interval of sensitivity of the extended definition of neglect was wide (54-74%) (**Table 3 and Table S4**). Furthermore, 47% of patients with neglect on the NIHSS had a missing or untestable prehospital assessment of either agnosia or tactile extinction (79/168).

## DISCUSSION

We found that the overall prehospital assessment of stroke code patients by paramedics correlates strongly with in-hospital examination by physicians, indicating that paramedics adequately assess stroke-related neurological deficits in the prehospital setting. Correlation was strongest in patients with a final diagnosis of aLVO ischemic stroke or intracranial hemorrhage. Concerning individual items, prehospital assessment of arm and leg motor function correlated best with in-hospital findings and had the highest sensitivity. In contrast, neglect, abnormal speech and gaze deviation had weakest correlation. Neglect and gaze deviation had lowest sensitivity, whilst abnormal speech had lowest specificity.

The most recent AHA/ASA stroke triage algorithm for ambulance paramedics recommends the use of a prehospital aLVO stroke scale.^1^ However, the quality of assessment of individual stroke-related deficits by paramedics was only studied in four small (n<190) studies and often in cohorts including only stroke patients rather than in unselected stroke code patients.^9-12^ A few other studies evaluated prehospital assessment of complete aLVO stroke scales by paramedics with assessment by physicians.^5-8^ One small study found very good agreement in assessment of the RACE scale by ambulance paramedics on-site and stroke neurologists using video telemedicine in 31 stroke code patients.^5^ Three other studies found that paramedics scored the Field Assessment Stroke Triage for Emergency Destination (FAST-ED) and Los Angeles Motor Scale (LAMS) similarly to emergency physicians and vascular neurologists.^6-8^ However, these studies had smaller sample sizes than our study and did not investigate separate clinical items. Additionally, the times between both assessments were generally longer than in our study, and one study compared LAMS assessment only in selected patients with a final diagnosis of stroke.^6^

Stratified by final diagnosis, we found that overall correlation was strongest for patients with an intracranial hemorrhage or aLVO ischemic stroke, who also had highest NIHSS scores. This indicates that the prehospital assessment by paramedics is especially adequate in case of severe neurological deficits, which is often the case in patients who require direct transportation to a TSC. Correlation was poorest in patients with a stroke mimic, possibly due to more prevalent fluctuations of symptoms in certain mimics, such as seizures or functional neurological disorders.

Our findings suggest that neglect is the most challenging item for paramedics to assess. In almost half of patients in whom neglect was observed in the hospital, this was missed at prehospital assessment. Furthermore, the positive predictive value of the prehospital assessment of neglect was very low. These findings are in line with a previous study in a small subgroup of patients with suspected stroke, in which neglect had the lowest inter-rater agreement between paramedics and the hospital stroke team.^10^ Following neglect, sensitivity was lowest for gaze deviation. Of note, these are both cortical symptoms and sensitive for aLVO ischemic stroke when assessed by an experienced neurologist, further emphasizing the importance of correct prehospital assessment of these symptoms for aLVO detection.^21^ Our findings suggest that in a setting with relatively well-trained paramedics the assessment of neurological deficits can still be enhanced, which might improve the value of the use of aLVO scales in prehospital stroke triage. Future research should address this, ideally also taking into account efficacy of other aLVO triage modalities, such as mobile stroke units, telemedicine, dry-electrode electroencephalography or transcranial doppler systems.^22^

Strengths of this study include the large sample size of unselected stroke code patients from multiple ambulance regions with several PSCs and TSCs. In addition, prehospital assessments were performed by paramedics in the field and implemented in regular workflow, representing the real-world setting and making results well generalizable for routine practice. Furthermore, due to the prospective designs, the amount of missing data was limited. This allowed for extensive evaluation of the separate clinical items, which enabled identification of the items that are most challenging for paramedics.

Limitations of this study include that subtle motor deficits could not be compared, since an arm and leg drift was scored as normal. This may in part explain the high sensitivity of motor function items, although correlation was still strongest for these items. Furthermore, we used the prehospital assessment of agnosia for comparison with neglect, whilst this does not encompass all modalities of the NIHSS definition of neglect. However, even in the additional analysis in the LPSS cohort with the addition of tactile extinction to agnosia, the sensitivity of neglect remained lowest. These results should be interpretated with caution since a high proportion of patients who had neglect on the NIHSS missed prehospital assessment of agnosia and tactile extinction. This again suggests that assessment of neglect is challenging, which is in line with previous literature.^10^ Possibly, since completing all items was not required in the LPSS, paramedics could have refrained from documenting prehospital assessment in certain cases, e.g. if patients could not be instructed because of a decreased consciousness or if patients were fixed for transport. We cannot exclude that this may have influenced the estimations of correlation, sensitivity and specificity. Furthermore, symptoms might have altered between the assessments due to spontaneous improvement or worsening, which may explain some of the disparities.^16-18^ However, we found no clear or clinically relevant trend in correlation after stratification for onset-to-ambulance-arrival time, the time interval between both assessments was relatively short (median 29 minutes) and patients with a final diagnosis of TIA were excluded. We therefore expect this effect to be limited. Also, even despite this possible disturbance, the overall correlation between both assessments was still strong. Furthermore, we would expect that a possible difference over time would apply to all prehospital items equally and therefore not explain differences between the individual items. Lastly, in the Netherlands, ambulance paramedics are well-trained registered nurses who received additional education in ambulance care. This may have improved their neurological examination and prehospital stroke assessment may be more challenging in regions where paramedics are less extensively trained.

## CONCLUSIONS

The overall prehospital assessment of stroke code patients by paramedics correlates strongly with in-hospital assessment by physicians, indicating that paramedics can adequately assess stroke-related neurological deficits in a prehospital setting. Prehospital assessment of neglect, abnormal speech and gaze deviation differed most from in-hospital assessment. Focused training on these three items may help to improve prehospital stroke triage.

### The LPSS and PRESTO collaborators

Leo A M Aerden, Kees C L Alblas, Jeannette Bakker, Eduard van Belle, Timo Bevelander, Jan Bosch, Bianca Buijck, Jasper D Daems, Luuk Dekker, Diederik W J Dippel, Tamara Dofferhoff-Vermeulen, Pieter Jan van Doormaal, Kirsten R I S Dorresteijn, Dion Duijndam, Martijne H C Duvekot, Roeland P J van Eijkelenburg, Adriaan C G M van Es, Jan-Hein Hensen, Amber Hoek, Henk Kerkhoff, Loet M H Kloos, Gaia T Koster, Nyika D Kruyt, Jan Willem Kuiper, Karlijn F de Laat, Arnoud M de Leeuw, Hester F Lingsma, Aad van der Lugt, Geert Lycklama À Nijeholt, Lisette Maasland, Bruno J M van Moll, Walid Moudrous, Laus J M M Mulder, Truc My Nguyen, Anja Noordam-Reijm, Erick Oskam, Aarnout Plaisier, Bob Roozenbeek, Anouk D Rozeman, Els L L M de Schryver, Esmee Venema, Marieke J H Wermer, Ruben M van de Wijdeven, Ido R van den Wijngaard, Annemarie D Wijnhoud, Merel L Willeboer, Mirjam Woudenberg, Mandy M A van der Zon, Erik W van Zwet, Egon D Zwets, Stas A Zylicz

## Data Availability

In compliance with Dutch law, patient data cannot be made available, since participants were not informed during the opt-out procedure about the public sharing of their individual participant data in de-identified form. The syntax and output files of the statistical analyses can be made available from the corresponding author upon reasonable request.

## Sources of Funding and Funding Statement

The LPSS was funded by the Dutch Brain Foundation (grant HA20 15.01.02), the Dutch Innovation Funds (grant 3.240), and Health-Holland (grant LSHM16041). The PRESTO study was funded by the BeterKeten collaboration and Theia Foundation (Zilveren Kruis). The funders had no role in study design, data collection, data analysis, data interpretation, or writing of the report, or the decision to submit for publication.

## Disclosures

MJHW reported receiving Clinical Established Investigator grant 2016T086 from the Dutch Heart Foundation and VIDI grant 9171337 from the Netherlands Organization for Health Research and Development (ZonMw) during the conduct of the original LPSS study. DWJD reports funding from the Dutch Heart Foundation, Brain Foundation Netherlands, The Netherlands Organisation for Health Research and Development, Health Holland Top Sector Life Sciences & Health, and unrestricted grants from Penumbra Inc., Stryker, Medtronic, Thrombolytic Science, LLC and Cerenovus for research, all paid to institution outside the submitted work. BR reported funding from the Dutch Heart Foundation and the Netherlands Organization for Health Research and Development (ZonMw) during the conduct of this study, paid to the institution. NDK reported receiving grant HA20 15.01.02 from the Dutch Brain Foundation, grant 3.240 from the Dutch Innovation Funds, and grant LSHM16041 from Health∼Holland during the conduct of the study. No other disclosures or conflicts of interests were reported.

## Non-standard Abbreviations and Acronyms

aLVO: anterior large-vessel occlusion
CI: confidence interval
EVT: endovascular thrombectomy
FAST: Face-Arm-Speech-Time
IVT: intravenous thrombolysis
IQR: interquartile range
LPSS: Leiden Prehospital Stroke Study
NIHSS: National Institutes of Health Stroke Scale
PSC: primary stroke center
PRESTO: PREhospital triage of patients with suspected STrOke study
RACE: Rapid Arterial oCclusion Evaluation
r_s_: Spearman’s rank correlation coefficient
SD: standard deviation
TIA: transient ischemic attack
TSC: thrombectomy-capable stroke center

## Notes

### Author Declarations

The protocols of the Leiden Prehospital Stroke Study and PREhospital triage of patients with suspected STrOke were approved by the relevant medical ethics review committees and institutional review boards of all participating centers.

